# Dual-Histamine Receptor Blockade with Cetirizine - Famotidine Reduces Pulmonary Symptoms in COVID-19 Patients

**DOI:** 10.1101/2020.06.30.20137752

**Authors:** Reed B. Hogan, Reed B. Hogan, Tim Cannon, Maria Rappai, John Studdard, Doug Paul, Thomas P. Dooley

## Abstract

**Background:** The COVID-19 pandemic due to SARS-CoV-2 infection can produce Acute Respiratory Distress Syndrome as a result of a pulmonary cytokine storm. Antihistamines are safe and effective treatments for reducing inflammation and cytokine release. Combinations of Histamine-1 and Histamine-2 receptor antagonists have been effective in urticaria, and might reduce the histamine-mediated pulmonary cytokine storm in COVID-19. Can a combination of Histamine-1 and Histamine-2 receptor blockers improve COVID-19 inpatient outcomes?

**Methods:** A physician-sponsored cohort study of cetirizine and famotidine was performed in hospitalized patients with severe to critical pulmonary symptoms. Pulmonologists led the inpatient care in a single medical center of 110 high-acuity patients that were treated with cetirizine 10 mg *b.i.d*. and famotidine 20 mg *b.i.d*. plus standard-of-care.

**Results:** Of all patients, including those with Do Not Resuscitate directives, receiving the dual-histamine receptor blockade for at least 48 hours, the combination drug treatment resulted in a 16.4% rate of intubation, a 7.3% rate of intubation after a minimum of 48 hours of treatment, a 15.5% rate of inpatient mortality, and 11.0 days duration of hospitalization. The drug combination exhibited beneficial reductions in inpatient mortality and symptom progression when compared to published reports of COVID-19 inpatients. Concomitant medications were assessed and hydroxychloroquine was correlated with worse outcomes.

**Conclusions:** This physician-sponsored cohort study of cetirizine and famotidine provides proof-of-concept of a safe and effective method to reduce the progression in symptom severity, presumably by minimizing the histamine-mediated cytokine storm. Further clinical studies in COVID-19 are warranted of the repurposed off-label combination of two historically-safe histamine receptor blockers.

## 1. Introduction

Histamine and mast cells play a fundamental role in modulating inflammation through increased capillary blood flow and vascular permeability, as well as cytokine release. Histamine-1 (H1) receptor antagonists (e.g., cetirizine) are administered for allergies. Histamine-2 (H2) receptor antagonists (e.g., famotidine) are used to control acid in the stomach and heart burn. Prescription branded, generic, and over-the-counter (OTC) drugs of both classes are safe and commercially available worldwide.

Humans have been treated using dual-histamine receptor blockade. Urticaria (hives) has been successfully treated with dual-histamine receptor blockade since the 1970’s [1-3], and remains a common practice in dermatology. And, a few reports have begun to demonstrate that diarrhea can be treated similarly [4, 5]. At present no H1-H2 receptor combination drug has been US FDA-approved.

COVID-19 (SARS-CoV-2) emerged in late 2019 in China, and nucleic acid sequence results indicate that it was very likely from a bat vector. The disease can manifest as a hyper-immune response with pulmonary cytokine release resembling that of other respiratory infections, such as Severe Acute Respiratory Syndrome (SARS), Middle East Respiratory Syndrome (MERS), and influenza. Studies from China have defined the COVID-19 cytokine profile [6] and identified risk factors that increase mortality [7]. These retrospective studies suggest mortality may be linked to inflammatory processes caused by a “cytokine storm”, which was very common in patients with severe to critical symptoms [8]. Pulmonary pathology in early-phase COVID-19 pneumonia has shown acute lung injury [9]. In the later stage of disease, patients can develop Acute Respiratory Distress Syndrome (ARDS) or ARDS-like conditions and multi-organ failure [10].

According to the Centers for Disease Control (CDC), the disease severity from China is 14% severe and 5% critical, and the critical patients displayed a fatality rate of 49%. (www.cdc.gov/coronavirus/2019-ncov/hcp/clinical-guidance-management-patients.html). Furthermore the CDC reports, “*Among U.S. COVID-19 cases with known disposition, the proportion of persons who were hospitalized was 19%. Among all hospitalized patients, a range of 26% to 32% of patients were admitted to the ICU. Among all patients, a range of 3% to 17% developed ARDS compared to a range of 20% to 42% for hospitalized patients and 67% to 85% for patients admitted to the ICU. Mortality among patients admitted to the ICU ranges from 39% to 72% depending on the study. The median length of hospitalization among survivors was 10 to 13 days.”*

Because of the sudden emergence of COVID-19, rapid research efforts are being conducted to repurpose existing approved drugs or biologic immunotherapies in this new indication, as these options are more likely to have near-term benefit during the pandemic. A major goal of many of these initiatives is to prevent or reduce the cytokine storm in pulmonary tissue [11-13].

Animal model studies are informative at this juncture. Sars-CoV-infected mice have shown that T-cell responses are required for protection from disease and for virus clearance [14]. The immunomodulation by histamine depends mostly on its influence of T-cells [15]. Histamine stimulates inflammation, cytokine release, and can lead to tissue damage, including lung [16]. A porcine study evaluating H1N1 influenza demonstrated the accumulation of histamine in severe pneumonia [17]. Furthermore, the H1 receptor antagonist, ketotifen, decreased inflammatory cytokines and severe pneumonia [17].

Dual-histamine receptor blockade has been effective in animal models of bacterial ARDS and allergy. A porcine animal model study has shown successful treatment of Pseudomonas-induced ARDS with diphenhydramine and cimetidine [18]. In this model for the treatment of hypoxemia, pulmonary hypertension, and pulmonary microvascular injury, the combination of diphenhydramine and cimetidine was essential, and was augmented somewhat by ibuprofen. In a guinea pig model, treatment with clemastine and cimetidine protected against allergen-induced bronchial obstruction [19].

Therefore, based upon prior efficacious dual-histamine receptor blocker studies in humans and animal models in a variety of diseases, we believe it is reasonable to bridge into humans infected by COVID-19 using dual-histamine receptor blockade, in order to prevent or diminish the cytokine storm. Furthermore, the safety and efficacy of dual-histamine receptor blockade previously evidenced in human urticaria (and diarrhea) makes this an appealing approach. Therefore, the major goals of the present proof-of-principle study include to decrease the progression of adverse outcomes in a cohort of severe and critical (high-acuity) hospitalized patients, most notably from ventilation independence to dependence, from life to death, and by reducing the duration of stay until discharge.

## 2. Methods

This physician-sponsored and -initiated cohort study was led by a team of board certified pulmonologists from a single practice group in Jackson, MS. All patients were treated in a single hospital operated by Baptist Health Systems, with IRB approval (IRB # 20-49 exemption) for retrospective access to patient data. The first dual drug-treated patient date was 3 April 2020 and the study period for this cohort concluded on 13 June 2020. The study was motivated by the principle of compassionate care use of repurposed medications (in view of the rationale above) by the attending pulmonologists during the COVID-19 pandemic. Conditions included open-label drug use, without a placebo control or randomization. In lieu of a placebo control group, for comparison the control(s) consisted of published SOC patient results that were not administered the dual-histamine receptor blockade from the USA, United Kingdom, and China.

### 2.1 Inclusion and Exclusion Criteria

The inclusion criteria were: (a) Males or females of minimum age of 17; (b) Admission to the hospital with suspected or confirmed pulmonary symptoms of COVID-19. All patients were confirmed COVID-19 positive by RT-PCR within several days of admission. The exclusion criteria were: (a) The patient is negative for COVID-19 by RT-PCR diagnostic test; (b) Sensitivity or allergy to cetirizine or famotidine, if known; (c) Duration of stay of less than 48 hours; and (d) Treatment with the drug combination for less than 48 hours. The investigators anticipated that any patient who was subject to a Do Not Resuscitate (DNR) directive may be a confounding factor (e.g., with regard to old age or the extent of aggressive life-sustaining care provided). Therefore, the DNR patients’ results were parsed in the analyses for comparison to all patients.

### 2.2 Standard-of-Care Procedures & Medications

On admission to hospital, the patient was diagnosed for suspected COVID-19 based primarily upon pulmonary symptoms, and located within a COVID-19 ward. Treatment was initiated in ER with SOC per admitting provider. The patient was confirmed positive for COVID-19 by RT-PCR diagnostic test. SOC included radiologic assessments, supplemental oxygen when necessary, and intravenous (IV) hydration when necessary. SOC concomitant treatments included the antimalarial drug hydroxychloroquine (84.5%), the anti-IL6 biologic tocilizumab (50.9%), the glucocorticoid drug methylprednisolone (30.9%), and convalescence plasma (30.0%). The cumulative rate of intubation in this cohort was 16.4%.

### 2.3 Cetirizine - Famotidine Treatment

The H1 receptor antagonist was cetirizine. The H2 receptor antagonist was famotidine. Given the current challenge of market availability of oral H2 antagonists, whenever the oral dosage form was not available or appropriate the clinicians used famotidine IV. Cetirizine and famotidine administration was preferably (a) oral, when feasible; then (b) gastric via nasogastric tube; then (c) by IV injection, based upon clinical assessments. The first dose of the therapy was administered in the ER, or upon arrival to the COVID ward, or when our trial started. Famotidine 20 mg IV and cetirizine 10 mg IV (or alternatively PO) was administered. Subsequent doses consisted of famotidine 20 mg q 12 hrs and cetirizine 10 mg q 12 hrs PO.

### 2.4 Study Endpoints

The major endpoints were: (a) Increased rate of discharge; (b) Reduced ventilation requirements (i.e., reduced number of intubations overall and after receiving a minimum of 48 hours of dual drug treatment); (c) Reduced inpatient mortality rate; and (d) Reduced duration of hospitalization.

The study endpoints were compared to SOC patient outcomes from Atlanta, GA [20], Louisiana [21], New York City, NY [22], United Kingdom [23], and Wuhan, China [24, 25]. This comparison to external sources is informative, as the number of available SOC-only patients was limited in this medical center during the rapidly-evolving SOC in the COVID-19 pandemic. This new dual drug treatment paradigm was rapidly adopted as SOC in this hospital.

## 3. Results

### 3.1 Patient Demographics

The patient demographics for the cetirizine - famotidine treatment cohort consisted of 110 patients age 17 to 97; mean age of 63.7 (SD 18.1); Female 59% and male 41%; and racial composition of 36.4% White, 59.1% African American or Black, 4.5% other. According to the US Census Bureau the racial demographics for the state of Mississippi are 59.1% White, 37.8% African American or Black, 3.1% other, and the median age is 36.7 years. Thus, our treated patients afflicted with COVID-19 in this medical center represented an inversion in racial demographics with regard to the state’s statistics vis-à-vis African-American or Black versus White.

### 3.2 Acuity and Comorbidity

The 110 COVID-19-positive patients with severe and critical pulmonary symptoms were treated in an inpatient setting in a single medical center with cetirizine and famotidine for a minimum of 48 hours, in addition to SOC. This group of patients manifested an average of 2.7 comorbidities (**Table I**). The most common comorbidities were hypertension (78.2%), obesity plus morbid obesity (58.2%), diabetes (42.7%), and cardiac disease (26.4%). Thus, the cohort exhibited high acuity and high comorbidities.

**Table I:**
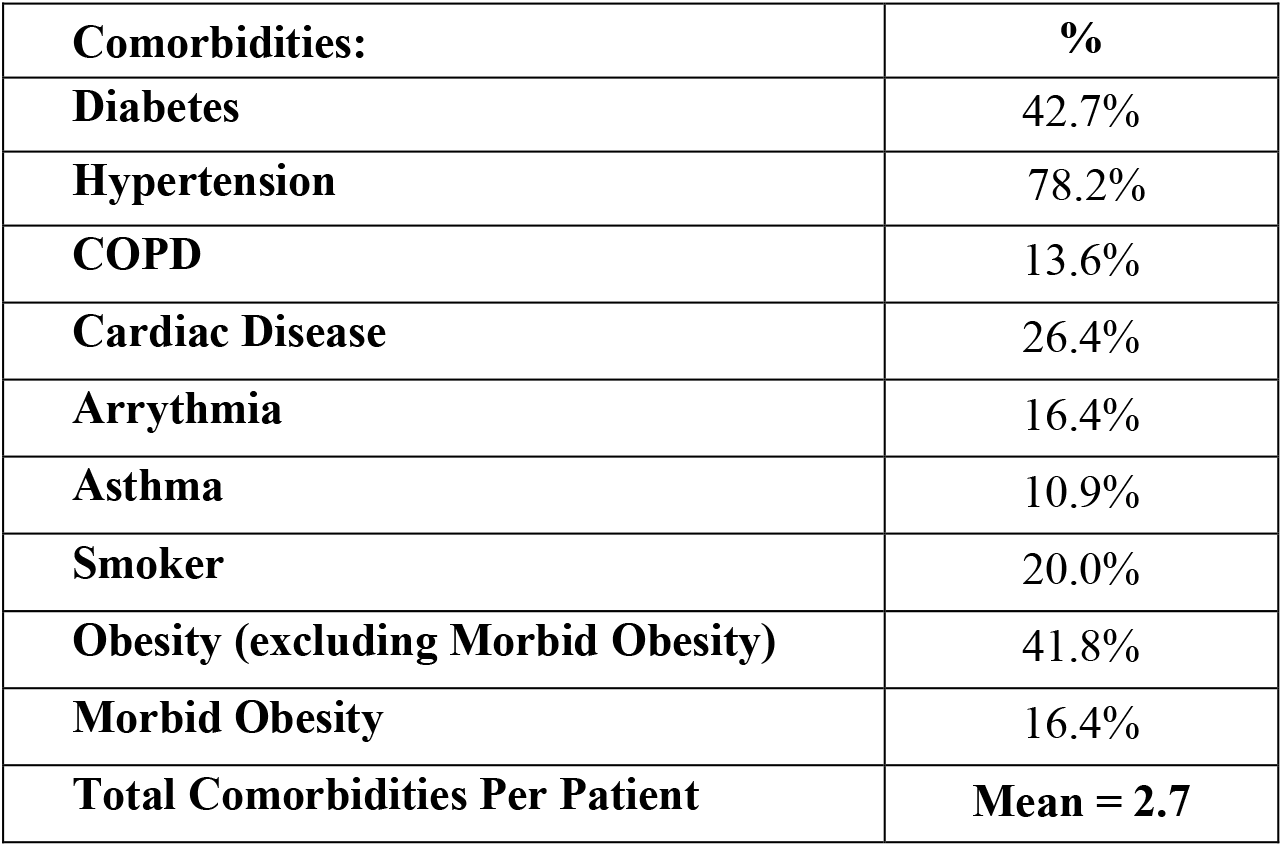
Comorbidities in 110 hospitalized COVID-19 patients treated with famotidine and cetirizine for a minimum of 48 hours.

| **Comorbidities:** | **%** |
| --- | --- |
| **Diabetes** | 42.7% |
| **Hypertension** | 78.2% |
| **COPD** | 13.6% |
| **Cardiac Disease** | 26.4% |
| **Arrhythmia** | 16.4% |
| **Asthma** | 10.9% |
| **Smoker** | 20.0% |
| **Obesity (excluding Morbid Obesity)** | 41.8% |
| **Morbid Obesity** | 16.4% |
| **Total Comorbidities Per Patient** | **Mean = 2.7** |

### 3.3 Clinical Results with and without Do Not Resuscitate Directives

The results of dual drug treatment are summarized with regard to all patients including DNR and the subset excluding DNR (**Table II**). The investigators anticipated DNR patients would be a confounding factor, and there were 13 DNR patients. The endpoints were: (a) discharge rate; (b) intubation rate; (c) intubation rate after a minimum of 48 hours of dual drug treatment; (d) inpatient mortality rate; and (e) average number of days to discharge. The rate of discharge during the study period (10 weeks plus 1 day) was 84.5% (and 91.8% excluding DNR). The intubation rate was 16.4% (and 16.5% excluding DNR). The intubation rate after a minimum of 48 hours of drug treatment was 7.3% (and 6.2% excluding DNR). The inpatient mortality rate was 15.5% (and 8.2% excluding DNR). The average number of days to discharge was 11.0 (and 10.9 days excluding DNR).

**Table II:** Clinical outcomes in 110 hospitalized COVID-19 patients treated with famotidine and cetirizine for a minimum of 48 hours.

| **Key Metric:** | **Including DNR** | **Excluding DNR** |
| --- | --- | --- |
| **Total Patients Admitted** | 110 | 97 |
| **Total Patients Discharged** | 93 | 89 |
| **Discharge Rate** | **84.5%** | **91.8%** |
| **Total Patients Intubated** | 18 | 16 |
| **Intubation Rate** | **16.4%** | **16.5%** |
| **Total Patients Intubated After a Minimum of 48 hrs of Treatment** | 8 | 6 |
| **Intubation Rate After a Minimum of 48 hrs of Treatment** | **7.3%** | **6.2%** |
| **Total Deaths** | 17 | 8 |
| **Death Rate %** | **15.5%** | **8.2%** |
| **Average Days to Discharge** | **11.0** | **10.9** |

### 3.4 Concomitant Medications, Treatments, and Intubations

Concomitant administration of other drugs, biologics, or treatments in the SOC of the severe to critical patients are potential confounding factors (**Table III**). For instance, the vast majority (84.5%) of the patients were administered hydroxychloroquine (HCQ). Based upon recent publications [26-29], one may assert that HCQ very likely provided no therapeutic benefit, and HCQ might have had an adverse effect on the outcomes in this cohort of 110 patients, as it correlated with worse outcomes (i.e., higher rates of intubation and death).

**Table III:** Clinical outcomes in 110 famotidine and cetirizine-treated COVID-19 patients and concomitant treatments.

| **Concomitant Treatments** | **Total #** | **% of Total** | **Intubated %** | **Death %** |
| --- | --- | --- | --- | --- |
| **Hydroxychloroquine - YES** | 93 | 84.5% | 18.3% | 17.2% |
| **Hydroxychloroquine - NO** | 17 | 15.5% | 5.9% | 5.9% |
| **Methylprednisolone - YES** | 34 | 30.9% | 29.4% | 14.7% |
| **Methylprednisolone - NO** | 76 | 69.1% | 10.5% | 15.8% |
| **Tocilizumab - YES** | 56 | 50.9% | 30.4% | 21.4% |
| **Tocilizumab - NO** | 54 | 49.1% | 1.9% | 9.3% |
| **Convalescent Plasma - YES** | 33 | 30.0% | 30.3% | 33.3% |
| **Convalescent Plasma - NO** | 77 | 70.0% | 10.4% | 7.8% |

In some of the patients, and especially those who progressed to ventilation dependence, a biologic (tocilizumab in 50.9% of patients), a glucocorticoid (methylprednisolone in 30.9% of patients), and/or convalescent plasma (in 30.0% of patients) were used at the discretion of the pulmonologist-led critical care team as concomitant treatments (**Table III**). At those junctures in patient management, the concomitant treatments were anticipated to possibly provide some benefit to the patients. However, the high degree of correlation of use of methylprednisolone, tocilizumab, and/or convalescent plasma with intubation per se hampered measurement.

It is interesting to note the clinical nature of the intubated patients and deaths associated with intubation. Of the 18 intubated patients, 9 died (50.0%). Of the group of 10 patients intubated *prior to* completing 48 hours of combination drug therapy, 9 were subsequently successfully extubated, 5 of whom were extubated within 2 to 4 days. Of the 8 patients intubated *after* completing a minimum of 48 hours of cetirizine and famotidine use, all 8 patients died and all received tocilizumab and convalescence plasma, whereas only 2 of the 8 patients received methylprednisolone. This particular patient subpopulation (n = 8) was critically ill and included 2 leukemia patients, a cirrhotic patient, a 91 year old with chronic kidney disease, a 79 year old former smoker with chronic kidney disease stage 3, an end stage renal disease patient, and a multiple sclerosis patient with chronic kidney disease in cardiogenic shock.

## 4. Discussion

Here we describe an initial study of dual-histamine receptor blockade to retard the histamine-cytokine network and with the intention of blunting the cytokine storm. The safety profile of dual-histamine receptor blockade makes this an appealing consideration for COVID-19-positive patients. If dual-histamine receptor blockade is able to blunt the cytokine system, the risk of progressing to severe and/or critical disease should lessen.

The results of this initial physician-sponsored study provide a proof-of-principle that it can reduce disease severity and the need for ventilators, and save lives. The results of patients who received at least 48 hours of the combination drug treatment demonstrated reduced rates of intubation (16.4%), of intubation after 48 hours of dual drug treatment (7.3%), of inpatient mortality (15.5%), and of duration of hospitalization (11.0 days). If DNR patients were excluded, then the inpatient mortality rate was only 8.2%. These clinical outcomes represent reductions in the anticipated symptom severity expressed as ventilator dependence and lethality relative to SOC reported in Atlanta, Georgia [20], Louisiana [21], New York City, New York [22], the United Kingdom [23], and Wuhan, China [24, 25] (see below).

### 4.1 Preliminary Results of Patients Not Treated with Cetirizine - Famotidine

Outside of the dual-histamine receptor blockade cohort group, preliminary results were noted by the same group of pulmonologists in the same medical center with an independent group of 12 SOC-only COVID-19 patients. The SOC-only patients (lacking cetirizine and famotidine) resulted in 5 intubations (41.7%), 5 deaths (41.7%), and 18.0 average days to discharge (data not shown). These SOC-only preliminary results are consistent with high symptom severity and high rate of inpatient fatality in the overall admitted patient population. Furthermore, a small group of 7 famotidine-only patients resulted in 3 intubations (42.9%), 1 death (14.3%), and 16.2 average days to discharge (data not shown). Due to the limited number of patients in each of these two groups, unfortunately they were not deemed sufficient for comparative statistical analysis, relative to 110 cohort patients receiving cetirizine and famotidine for a minimum of 48 hours.

### 4.2 Inpatient Fatality Rate

The 15.5% inpatient fatality rate or 8.2% excluding DNR patients compares favorably to published inpatient fatality rates from other regions -- 25.8% in Atlanta, GA [20], 23.6% in Louisiana [21], 21% in New York City, NY [22], 25.7% in the large RECOVERY trial in the United Kingdom [23], and 21.9 and 28.3% in Whuan, China [24, 25]. In other words, we experienced a reduction in inpatient fatalities of approximately one fourth to one third relative to these reference locations ranging from 21% to 28.3% in inpatient fatality rates, some of which were obtained from very large clinical trials.

The reports of inpatient mortality rate can be dependent on multiple variables, such as inclusion/exclusion criteria (e.g., DNR patients), diagnosis (e.g., presumptive COVID-19 vs. PCR-confirmed for viral RNA), demographics of patients in the study, the duration of the study period, symptom severity and comorbidities at the time of admission, as well as rapidly evolving SOC treatments influenced by media, governmental agencies, and clinical reports. During this crisis it should be noted that many of the “publications” on COVID-19 were available only in preprint form, in view of time-is-of-the-essence. And, in some instances the information was only presented as an assertion in the media, without any supporting scientific information.

That being said, in our cohort 17 fatalities had an average age of 70.6 and our 9 DNR deaths had an average of 75.8 years. *(Note that one DNR death was due to a cirrhotic patient aged 38. If this individual had been excluded, then the DNR deaths would have averaged 80.5 years.)* Our overall and DNR deaths were predominantly among the elderly. Furthermore, the small group of 12 SOC-only patients treated by our pulmonologists exhibited a high inpatient fatality rate of 41.7% in the same medical center (data not shown). These two findings (i.e., elderly patient deaths and a small SOC-only group with a very high rate of inpatient fatality), suggest that our inpatient fatality rate of 15.5% with DNR was adversely impacted by patient age and multiple comorbidities (average of 2.7) in Central Mississippi. In other words, our dual-histamine receptor blockade treatment effects were favorable even in an unfavorable context in high acuity patients.

### 4.3 Ventilator Dependence

With regard to ventilator dependence, the observed intubation rate of 16.4% overall in the cohort of 110 patients (including DNR directives) in Jackson, Mississippi and especially only 7.3% intubation rate after a minimum of 48 hours of treatment with cetirizine plus famotidine compares favorably to 26.3% in Louisiana [21], 12.2% in New York City [22], and 33.3% in Wuhan, China [24].

### 4.4 Limitations

There are multiple limitations to any physician-sponsored cohort (or case series) study. Within this initial study the limitations were most notably: (a) This proof-of-principle study was not a placebo-controlled, randomized, and blinded study that is customary for a regulatory registration trial; and (b) The study lacked a sufficiently high number of untreated SOC patients for use as a retrospective control cohort. However, the investigators have provided comparisons to the published SOC cohorts from other regions in the USA, UK, and China.

However, offsetting these limitations it should be noted that this work was performed in April through June 2020 during an intense season within the COVID-19 pandemic crisis, at a time-is-of-the-essence season when pulmonologists, emergency room physicians, critical care specialists, and hospitalists were eager to attempt rational repurposing of previously FDA-approved medications. Thus, the physicians desired near-term improved outcomes, compassionate care, and to identify new proof-of-concept off-label therapies for COVID-19 ARDS or ARDS-like patients. Cetirizine and famotidine were intentionally incorporated into treatment of this severe and critical patient cohort in view of exceptional historic safety of each medication, as evidenced by their approved OTC status, and in view of prior efficacy of dual-histamine receptor blockade in diseases in humans and animal models. This resulted in the vast majority of all COVID-19 positive patients in this medical center being treated with cetirizine plus famotidine as the “new” SOC.

### 4.5 Current Trends in Experimental COVID-19 Treatments

How does this H1-H2 drug combination treatment compare with other recent therapeutic developments in COVID-19? There are four noteworthy examples that have received attention at this juncture in the scientific literature and media, namely hydroxychloroquine, remdesivir, famotidine, and dexamethasone.

### 4.6 Hydroxychloroquine

First, hydroxychloroquine (HCQ) rapidly received general acceptance as a SOC medication for COVID-19 inpatients, and was in common use in critical care settings in the USA at the time of our study. The risk-benefit reward analysis of HCQ rapidly evolved to a highly unfavorable impression, based upon controlled clinical trials. The efficacy of HCQ in COVID-19 is now seriously doubted, and at least one cardio-toxic side effect has been noted [26]. This study consisted of 75 patients per arm, comparing SOC vs SOC + HCQ. The authors concluded that HCQ was not beneficial and resulted in more adverse events.

In another study HCQ was also ineffective in inpatients who required oxygen; 84 patients were treated with HCQ within 48 hours of admission vs 97 patients receiving SOC without HCQ [28]. There were no benefits to HCQ when assessing: (a) survival without transfer to ICU; (b) survival at 21 days; (c) survival without ARDS at 21 days; and (d) percentage of patients requiring oxygen at 21 days. Furthermore, 10% of the HCQ-treated patients manifested ECG anomalies requiring discontinuation of HCQ treatment. HCQ is no longer recommended for use in COVID-19 inpatients (i.e., *late* in disease progression), and the US FDA rescinded the Emergency Use Authorization (EUA) for this drug.

Two prospective well controlled clinical trials are high level evidence of a lack of efficacy of HCQ in *early* disease prevention or treatment in COVID-19 outpatients. A randomized, double-blind, placebo-controlled treatment trial with 423 outpatients concluded, “*Hydroxychloroquine did not substantially reduce symptom severity in outpatients with early, mild COVID-19.”* Side effects were greater with HCQ than placebo [29]. Another randomized, double-blind, placebo-controlled prevention trial with 719 individuals with high-risk exposure to a confirmed COVID-19 contact did not prevent illness when used as post-exposure prophylaxis within 4 days after exposure [27]. Side effects were greater with HCQ than placebo [27]. However, in spite of these solid clinical trial results, at this juncture many anecdotes about HCQ are being widely reported in the media, while lacking published scientific validation. Whether nuanced dosing, scheduling, and/or coincident medical treatments with HCQ might be efficacious in COVID-19 outpatients is an open question at this juncture.

Regardless, in aggregate these studies indicate that HCQ was not effective in preventing or treating disease progression in COVID-19 patients. Therefore, it should be noted that HCQ was administered to most of the patients in our study, and it is reasonable to speculate that it impaired patients in our cohort, in view of the demonstrated correlation with worse outcomes.

### 4.7 Remdesivir

Second, remdesivir was developed prior to the COVID-19 pandemic as a retroviral replication inhibitor. A well-designed placebo-controlled trial suggested a reduction in the time to clinical improvement [30]. However, the study was not sufficiently powered statistically [31]. Thus, there was no improvement with regard to patient deaths or viral load. This trial did not provide convincing evidence of a substantial therapeutic benefit of remdesivir. Regardless, the US FDA granted EUA for this prescription medication, which is expected to be expensive in the USA.

### 4.8 Famotidine

Third, famotidine has been evaluated in a retrospective association study of COVID-19 patients in New York City [32]. A cohort of 84 hospitalized patients out of 1,620 total received famotidine within 24 hours of hospitalization, and a subset of the patients (15%) were already treated with famotidine at home prior to admission. Patients intubated within 48 hours of admission were excluded. The doses ranged from 10-40 mg. Famotidine use was associated with a reduced risk of death and intubation. By comparison proton pump inhibitors (that reduce gastric acid independent of a histamine mechanism) were not associated with reduced risk of death or intubation. The results suggest a histamine-mediated effect in the COVID-19 patients. This report lacks information on concomitant medications or treatments (either SOC or experimental), with the exception of proton pump inhibitors [32].

Comparing our cetirizine + famotidine results with high comorbidity patients to the famotidine-only study [32], note the following comorbidity and other factors might account for the magnitude of treatment effect in our 110 cetirizine + famotidine patients vs. 84 famotidine patients in New York City: obesity 58.2% vs. 26%; hypertension 78.2% vs. 35%; diabetes 42.7% vs. 29%. In addition, the representation of Black patients was 59.1% vs. 21%. These comorbidity and racial factors are known to be causal in adverse COVID-19 inpatient outcomes. In addition, the overall intubation rate in all patients in New York City was only 8.8% and excluded patients intubated within 48 hours of admission. Once again this indicates a lower relative acuity level in the famotidine-only cohort of 84 patients. Our cohort’s intubation rate was 16.4% overall, whereas after a minimum of 48 hours of cetirizine + famotidine treatment it was only 7.3%. In addition, the inpatient mortality rate in all 1620 patients was 15% in the New York City study, which was essentially a lower baseline than expected for our Mississippi cohort.

These acuity, comorbidity, and outcome differences underscore a very likely comorbid burden in our patient cohort in Mississippi, relative to the famotidine-only study in New York City. Furthermore, concomitant medications (or other treatments) were not reported in the New York City study, whereas concomitant medicines were reported herein in **Table III** regarding HCQ (84.5%), methylprednisolone (30.9%), tocilizumab (50.9%), and convalescent plasma (30.0%). Note that HCQ correlated with worse outcomes in our study, and likely further burdened our patients.

In another retrospective inpatient study of 878 COVID-19 patients total, 83 patients were treated with famotidine alone in Hartford, Connecticut (Mather J.F., et al., Am. J. Gastroenterology, 2020, in press). In all patients the inpatient fatality rate was 21.8% and ventilation rate was 27.2%. Famotidine was administered at 20 mg/day or 40 mg/day, and resulted in an inpatient fatality rate of 14.5% and intubation rate of 21.7%.

Similar to what is described above for the New York City study, several key comorbidity factors were more prevalent in our study of cetirizine plus famotidine vs. the Hartford study: obesity 58.2% vs. 4.8%; hypertension 78.2% vs. 32.5%; diabetes 42.7% vs. 21.7%. These large differences in underlying medical conditions favored the health of the Hartford cohort of 83 patients. Furthermore, in a matched comparison of 83 famotidine patients vs. a subgroup of 689 no famotidine patients, HCQ was administered less frequently in the famotidine patients (43.4% famotidine vs. 52.0% no famotidine). HCQ is expected to have an adverse effect in hospitalized patients. And, corticosteroids were administered more frequently in the famotidine patients (57.8% famotidine vs. 47.8% no famotidine). Corticosteroids are likely to have a beneficial effect in hospitalized patients (see “dexamethasone” below). Thus, it is possible that these two concomitant medications biased the famotidine-treated group favorably and disadvantaged the no famotidine control group to some extent.

Taken together three studies with moderately sized treatment cohorts (range of 83 to 110 patients) that were administered cetirizine plus famotidine (this work) or famotidine-alone [32] (and Mather et al, in press) have shown efficacy in COVID-19 inpatients. Controlled prospective trials that are randomized with equivalent levels of acuity, comorbidity, concomitant medications, and other determinants (e.g., race) would be welcome to assess the relative contributions of famotidine and/or cetirizine in the presumptive blocking of the cytokine storm in COVID-19.

Although less informative than the two moderately sized cohort studies in New York City and Hartford, a case series of only 10 outpatients administered very high dose famotidine (most of them received 80 mg t.i.d. = 240 mg daily) perhaps suggested the possibility of a benefit [33].

For comparison to the three famotidine-only studies, our inpatient study used 20 mg twice daily (40 mg daily) of famotidine, which is within the FDA-approved OTC dosage level. However, the cetirizine amount was 10 mg twice daily in this inpatient context, which is double the daily FDA-approved dosage as an OTC medication.

### 4.9 Dexamethasone

The RECOVERY trial in the United Kingdom is a randomized, controlled, open-label trial of hospitalized patients [23]. Dexamethasone, a glucocorticoid, was administered to 2104 patients compared to 4321 patients receiving SOC. The steroid treatment reduced the 28-day fatality rate down to 22.9% vs. 25.7% in the SOC control patients. Also, dexamethasone reduced the incidence of death of patients on mechanical ventilation down to 29.3% compared to 41.4% in the SOC control patients.

In aggregate these controlled trials and cohort studies reveal at this juncture that COVID-19 inpatients in general are likely to benefit from: (a) cetirizine - famotidine combination or famotidine alone; (b) remdesivir; and/or (c) dexamethasone; but, not from hydroxychloroquine.

## 5. Conclusions

What is the current context and unmet medical need for repurposing old active ingredients, especially OTC medications, during the COVID-19 Pandemic? Given (a) the recent emergence of COVID-19, (b) the rapid need for safe and effective treatments deployable immediately, and (c) the rapid evolution in the SOC treatments for inpatients, recent innovations might not permit sufficient time for the statistically robust clinical trials that are customary for an FDA regulatory approval process. In this time-is-of-the-essence pandemic context, the results of this dual-histamine receptor blockade treatment compares favorably to current SOC protocols. We have demonstrated reductions in ventilator dependence and inpatient fatality rates relative to other published studies using two safe OTC medications. This dual drug approach is consistent with the historic axiom in medicine - “first do no harm”, by utilizing historically well tolerated OTC medications.

Although this study provides initial evidence in support of a safe and effective treatment, many questions remain to be addressed. Randomized and controlled trials are warranted with outpatients “early” and inpatients “late”. We have initiated a separate randomized placebo-controlled outpatient trial of cetirizine - famotidine to begin to address this question in PCR-confirmed asymptomatic or mild-to-moderate severity outpatients, thus early in disease progression. Furthermore, is the beneficial effect dependent on this particular combination of active pharmaceutical ingredients, or would alternative H1 and/or H2 receptor antagonists also be effective? Given the historic safety profiles for the two selected ingredients within approved OTC monotherapies, it follows that one is likely to prefer these two over other active ingredients that have not been granted OTC status. And, would some patients benefit (more) from alternative doses or dosage schedules?

Given that famotidine alone has been reported to be beneficial in COVID-19 inpatients, it would be desirable to conduct a factorial-design randomized trial with cohorts having equivalent (a) acuities, (b) comorbidities, (c) concomitant medications, and (d) appropriate dosing to address the relative contributions of cetirizine and famotidine. It would also be beneficial to exclude hydroxychloroquine as a confounding factor in prospective clinical trials, as it appears to have been ineffective and/or detrimental in other retrospective inpatient trials, as well as in our cohort of high acuity patients.

The present clinical investigation provides a new method of treatment for this unmet medical need. The favorable circumstances of having commercially-available branded, generic, and OTC drugs targeting both H1 and H2 receptor types provide another distinct advantage relative to other experimental drug and biologic research programs in COVID-19, some of which may take numerous years to develop and commercialize. This approach with OTC cetirizine (e.g., Zyrtec®) and OTC famotidine (e.g., Pepcid®) could be rapidly deployed worldwide and should be affordable, even for economically under-served populations, not just for the economically advantaged.

## Data Availability

The 110 patient cohort data spreadsheet will be made available to the public upon acceptance of the manuscript. An Excel spreadsheet file will be available.
Reference regarding guidance for cohort or case series medical publications:
"Appropriate Use and Reporting of Uncontrolled Case Series in the Medical Literature." J.H. Kempen, (2011) Am J Ophthalmol. 2001;151(1): 7-10.el. doi 10.1016/j.ajo.2010.08.047

## Acknowledgements

The authors gratefully acknowledge the assistance of the clinical staff and administrators of Baptist Health Systems (including Dr. Mike Byers) and Jackson Pulmonary Associates. A preprint was posted online after manuscript submission: https://www.medrxiv.org/content/10.1101/2020.06.30.20137752v1

## Author Contributions

Dr. Hogan II - conceptualization, methodology, project administration, supervision, writing - review; Dr. Hogan III - conceptualization, methodology, project administration; Drs. Cannon, Rappai, and Studdard - conceptualization, data curation, investigation, methodology, project administration, writing - review; Dr. Paul - data curation, formal analysis, validation, visualization, writing - review; Dr. Dooley - methodology, formal analysis, validation, visualization, writing - original draft, revised draft.

## Supplementary Material

The data set of clinical parameters of the 110 inpatient cohort is provided, linked to this article (______).

## References

1. Wan, K.S., Efficacy of leukotriene receptor antagonist with an anti-H1 receptor antagonist for treatment of chronic idiopathic urticaria. J Dermatolog Treat, 2009. 20(4): p. 194–7.

2. Phanuphak, P., A. Schocket, and P.F. Kohler, Treatment of chronic idiopathic urticaria with combined H1 and H2 blockers. Clin Allergy, 1978. 8(5): p. 429–33.

3. Monroe, E.W., et al., Combined H1 and H2 antihistamine therapy in chronic urticaria. Arch Dermatol, 1981. 117(7): p. 404–7.

4. Hassoun, Y., M.R. Stevenson, and D.I. Bernstein, Idiopathic postprandial diarrhea responsive to antihistamines. Ann Allergy Asthma Immunol, 2019. 123(4): p. 407–409.

5. Mohammadi, E., et al., 287 - Exploring an Antihistamine Combination Therapy for Diarrhea Predominant Irritable Bowel Syndrome. Gastroenterology, 2018. 154(6): p. S–72.

6. Huang, C., et al., Clinical features of patients infected with 2019 novel coronavirus in Wuhan, China. The Lancet, 2020. 395(10223): p. 497–506.

7. Ruan, Q., et al., Clinical predictors of mortality due to COVID-19 based on an analysis of data of 150 patients from Wuhan, China. Intensive Care Medicine, 2020. 46(5): p. 846–848.

8. Mehta, P., et al., COVID-19: consider cytokine storm syndromes and immunosuppression. The Lancet, 2020. 395(10229): p. 1033–1034.

9. Tian, S., et al., Pulmonary Pathology of Early-Phase 2019 Novel Coronavirus (COVID-19) Pneumonia in Two Patients With Lung Cancer. Journal of Thoracic Oncology, 2020. 15(5): p. 700–704.

10. Cao, Y., et al., Imaging and clinical features of patients with 2019 novel coronavirus SARS-CoV-2: A systematic review and meta-analysis. Journal of Medical Virology, 2020. n/a(n/a).

11. Maude, S.L., et al., Managing cytokine release syndrome associated with novel T cell-engaging therapies. Cancer journal (Sudbury, Mass.), 2014. 20(2): p. 119–122.

12. Conti, P., et al., Induction of pro-inflammatory cytokines (IL-1 and IL-6) and lung inflammation by Coronavirus-19 (COVI-19 or SARS-CoV-2): anti-inflammatory strategies. J Biol Regul Homeost Agents, 2020. 34(2).

13. Zhang, W., et al., The use of anti-inflammatory drugs in the treatment of people with severe coronavirus disease 2019 (COVID-19): The Perspectives of clinical immunologists from China. Clin Immunol, 2020. 214: p. 108393.

14. Zhao, J., J. Zhao, and S. Perlman, T cell responses are required for protection from clinical disease and for virus clearance in severe acute respiratory syndrome coronavirus-infected mice. J Virol, 2010. 84(18): p. 9318–25.

15. Kmiecik, T., et al., T lymphocytes as a target of histamine action. Arch Med Sci, 2012. 8(1): p. 154–61.

16. Branco, A., et al., Role of Histamine in Modulating the Immune Response and Inflammation. Mediators Inflamm, 2018. 2018: p. 9524075.

17. Jang, Y., M. Jin, and S.H. Seo, Histamine contributes to severe pneumonia in pigs infected with 2009 pandemic H1N1 influenza virus. Arch Virol, 2018. 163(11): p. 3015–3022.

18. Sielaff, T.D., et al., Successful treatment of adult respiratory distress syndrome by histamine and prostaglandin blockade in a porcine Pseudomonas model. Surgery, 1987. 102(2): p. 350–7.

19. Dorsch, W., H.J. Reimann, and J. Neuhauser, Histamine1--histamine2 antagonism: effect of combined clemastine and cimetidine pretreatment on allergen and histamine-induced reactions of the guinea pig lung in vivo and in vitro. Agents Actions, 1982. 12(1-2): p. 113–8.

20. Auld, S., et al., ICU and ventilator mortality among critically ill adults with COVID-19. medRxiv, 2020: p. 2020.04.23.20076737.

21. Price-Haywood, E.G., et al., Hospitalization and Mortality among Black Patients and White Patients with Covid-19. N Engl J Med, 2020. 382(26): p. 2534–2543.

22. Richardson, S., et al., Presenting Characteristics, Comorbidities, and Outcomes Among 5700 Patients Hospitalized With COVID-19 in the New York City Area. JAMA, 2020.

23. Group, R.C., et al., Dexamethasone in Hospitalized Patients with Covid-19 - Preliminary Report. N Engl J Med, 2020.

24. Wu, C., et al., Risk Factors Associated With Acute Respiratory Distress Syndrome and Death in Patients With Coronavirus Disease 2019 Pneumonia in Wuhan, China. JAMA Intern Med, 2020.

25. Zhou, F., et al., Clinical course and risk factors for mortality of adult inpatients with COVID-19 in Wuhan, China: a retrospective cohort study. Lancet, 2020. 395(10229): p. 1054–1062.

26. Tang, W., et al., Hydroxychloroquine in patients with mainly mild to moderate coronavirus disease 2019: open label, randomised controlled trial. BMJ, 2020. 369: p. m1849.

27. Boulware, D.R., et al., A Randomized Trial of Hydroxychloroquine as Postexposure Prophylaxis for Covid-19. N Engl J Med, 2020.

28. Mahevas, M., et al., Clinical efficacy of hydroxychloroquine in patients with covid-19 pneumonia who require oxygen: observational comparative study using routine care data. BMJ, 2020. 369: p. m1844.

29. Skipper, C.P., et al., Hydroxychloroquine in Nonhospitalized Adults With Early COVID-19: A Randomized Trial. Ann Intern Med, 2020.

30. Wang, Y., et al., Remdesivir in adults with severe COVID-19: a randomised, double-blind, placebo-controlled, multicentre trial. Lancet, 2020. 395(10236): p. 1569–1578.

31. Norrie, J.D., Remdesivir for COVID-19: challenges of underpowered studies. Lancet, 2020. 395(10236): p. 1525–1527.

32. Freedberg, D.E., et al., Famotidine Use is Associated with Improved Clinical Outcomes in Hospitalized COVID-19 Patients: A Propensity Score Matched Retrospective Cohort Study. Gastroenterology, 2020.

33. Janowitz, T., et al., Famotidine use and quantitative symptom tracking for COVID-19 in non-hospitalised patients: a case series. Gut, 2020.

